# Violin plot data: A concerto of crucial information on valve thrombogenicity classified using laboratory measured valve motion

**DOI:** 10.1101/19009407

**Authors:** Lawrence N. Scotten, David J. Blundon, Marcus-André Deutsch, Rolland Siegel

**Author notes:** **Correspondence:** Lawrence N. SCOTTEN, 1560 Bonair Place, Victoria, BC, V8P 4V4, Canada. **Sources of funding:** This research did not receive any specific grant from funding agencies in the public, commercial, or not-for-profit sectors.

## Abstract

**Graphical Abstract:** 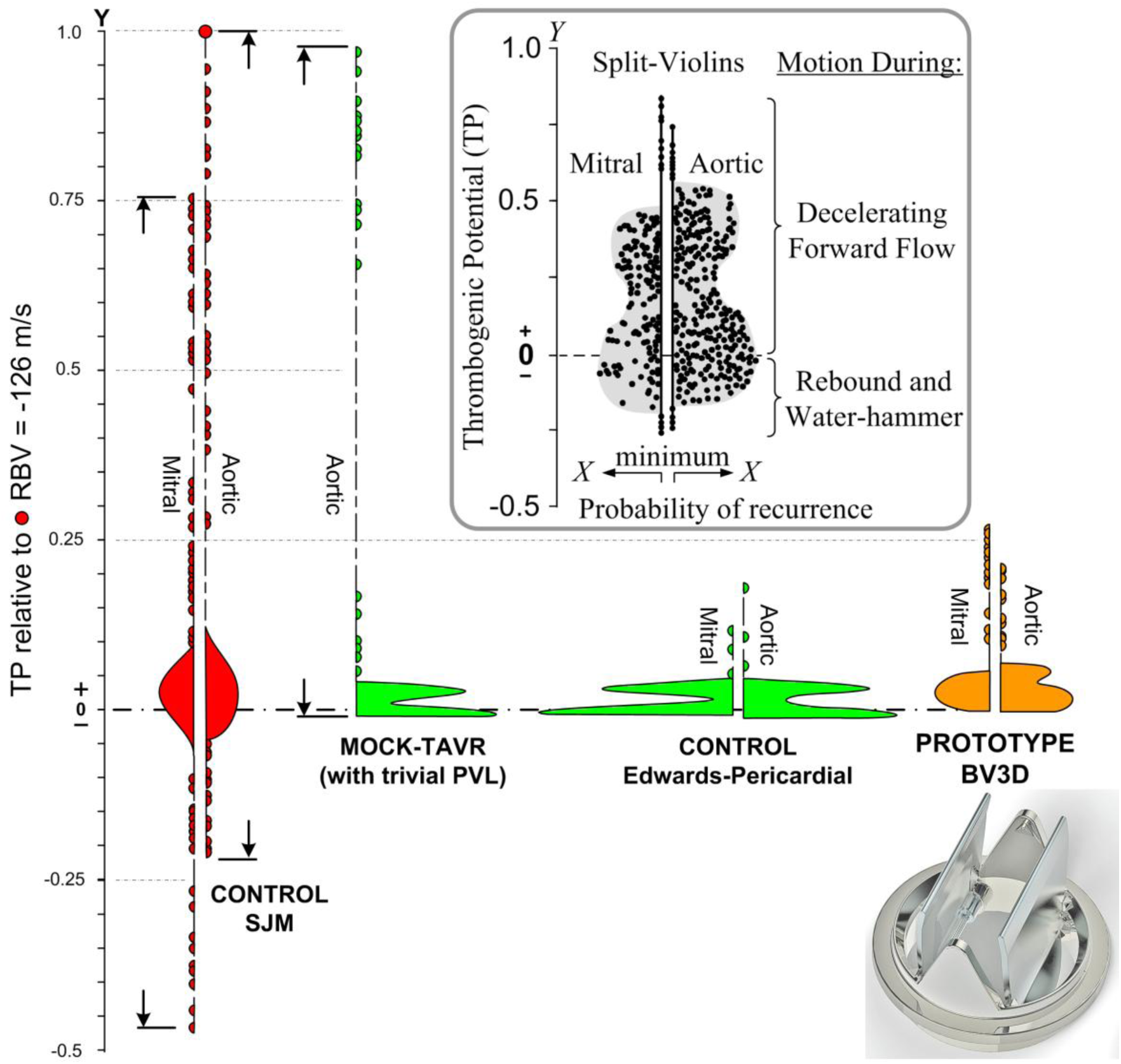

**Structured Abstract:** *Background:* A thrombogenic potential metric was used to compare prosthetic valve models in vitro.

*Methods:* Valves were tested in aortic and mitral sites under pulsatile circulation in a pulse duplicator. An optical approach measures dynamic valve area. Pulsatile fluid dynamics were measured by conventional techniques and a quasi-steady flow tester was used to measure valve leakage. Regurgitant flow velocity was derived using time-dependent volumetric flow rate / dynamic valve area. Since flow velocity and fluid shear force are related through flow velocity gradient, thrombogenic potential for valves that achieve near closure during the forward flow deceleration phase were determined as regurgitant flow velocities relative to the control mechanical valve regurgitant flow velocity of −126 m/s. Analysis of the flow velocity data made use of Vioplot R* and VISIO software packages to provide split violin plots that represent probability of recurrence of data.

*Results:* Thrombogenic potential was made dimensionless and ranged between −0.45 and +1.0. Negative thrombogenic potentials arise when transient rebound of valve occluder is accompanied by water-hammer phenomena. Positive thrombogenic potentials occur during decelerating forward flow. Bioprostheses had lowest thrombogenic potential transient of 0.15. A mock-transcatheter aortic valve replacement incorporating by design a trivial paravalvular leak (∼1.35 ml/s) demonstrated a high transient thrombogenic potential of 0.95.

*Conclusions:* Our data reveals distinct thrombogenic potential profile differences between valve models. If our study methods are verifiable, the design of future valves may utilize currently available experimental tools to produce advanced devices with significantly reduced thrombogenic potential.

*Highlights:*

➢ Prosthetic valve shortcomings include chronic unresolved thrombogenicity and durability issues.
➢ Thrombogenic potential of mechanical valves is fueled by brief, high closing flow velocities.
➢ Trivial leakage in bioprosthetic valves can yield high thrombogenic potential.
➢ Valve motion and flow velocity metrics may help advance prosthetic valve designs.

## 1. Introduction and Background

Nearly 60 years after their introduction, modifications to mechanical and bioprosthetic valves have not yet resolved chronic shortcomings related to durability and thrombogenic potential. Early published data on sources of thrombogenicity in mechanical valves by Davey [1] in 1966, visualized cyclic flow behavior by high-speed cinematography in a pulse duplicator with a transparent chamber and a sheet of light flow technique using aluminum flow tracer particles in transparent test fluids. Flow patterns, adjacent to an early model Starr-Edwards valve ball-occluder, were identified and multiple retrograde impulses were observed at valve closure. Although the investigators speculated that these events were associated with high shear, equipment of that era was incapable of measuring the retrograde flow velocities associated with occluder rebound phenomena [Davey, TB]. More recently, computational methods employed to examine flow patterns, flow velocities, and pressures associated with contemporary mechanical and bioprosthetic valve function have generated extensive information on flow but as yet have not proposed a development pathway to eliminate thrombogenicity in the former and extend durability in the latter [e.g. 2-4].

In prior publications, [5-9] we hypothesized that mechanical valve occluder closing behavior was implicated in the thrombogenic discrepancy between mechanical and bioprosthetic valves. Our modified pulse duplicator allowed measurement of prosthetic valve dynamic area (PDVA) providing crucial insight into occluder forward flow flutter, cavitation and occluder rebound that contribute to the complex genesis of valve thrombosis. Subsequent pulse duplicator adaptations included a unique electro-optical subsystem referred to as Leonardo [6] that revealed spatially averaged flow velocity in immediate proximity to test valves. We found that, transient regurgitant flow velocities (RFVs), occurred often near MHV closure, and proposed it as a proxy [6-9] for rapidly changing shear forces. Shear forces are known to promote formation of micro-thrombi aggregates that may remain locally adherent or embolize. This work provided incentive to refine geometry of a mechanical heart valve (MHV) with blood damage potential reduced to tissue valve levels. Included in these investigations were modified bileaflet [7] and trileaflet designs [6, 10, 11]. Also, this experimental methodology was extended to study thrombogenic potential in transcatheter aortic valve replacement (TAVR) devices [8, 9].

## 2. Methods

### 2.1 Test Apparatus

An extensively cited commercial in vitro test system [3, 5-9] was used with typical simulation conditions including pulse rate 70 cycles/min; pressures ca.120/80 mmHg (millimeters of mercury); and cardiac output 5 litres/min. Basic test methodology is depicted in Figure 1.

**Figure 1:**
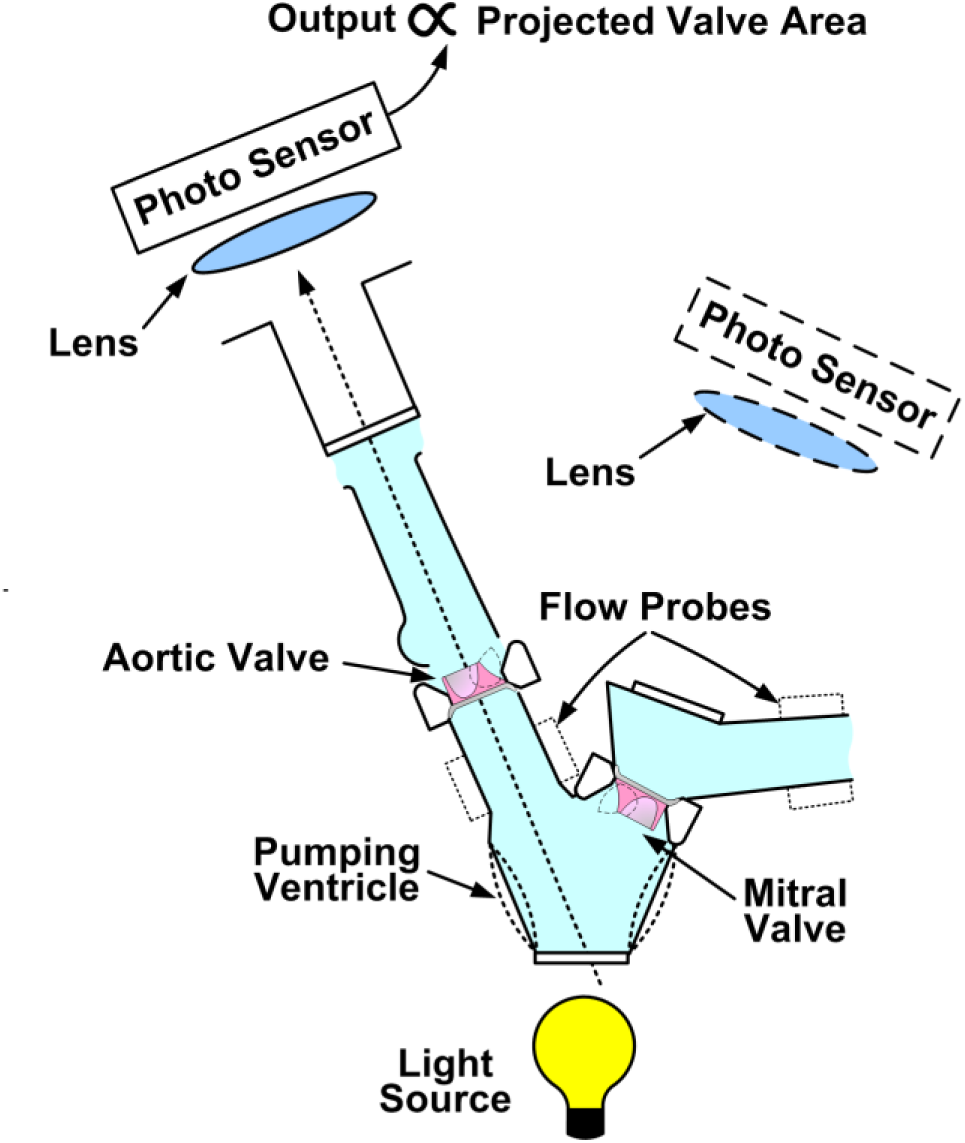
Basic test methodology uses optical approach to gauge aortic and mitral prosthetic heart valve motion quantified as projected dynamic valve area (PDVA).

Test valves included: a CLINICAL MECHANICAL (SJM, St Jude Medical Regent^TM^; Abbott Laboratories, Abbott Park, Ill, USA); a RAPID PROTOTYPE MECHANICAL (BV3D) [7]; a CONTROL BIOPROSTHESIS

Carpentier-Edwards Perimount (Edwards Lifesciences, Irvine, CA, USA) and a MOCK-TAVR construct fabricated using a bovine pericardial aortic valve Carpentier-Edwards Perimount mounted in a purpose-built fixture [8].

Nominal valve sizes were 25 mm tissue annulus diameter (TAD). Valves tested in pulsatile and quasi-steady pressure/flow systems and PDVA in pulsatile flow was obtained. Time-dependent inferred RFV was derived by dividing the volumetric flow rate by the PDVA (volumetric flow rate/PDVA). A separate leakage tester (ViVitro Labs Inc., model LT8991, circa 1998) provided accurate measurements of small closed valve leakage rates, under quasi-steady gravity head generated pressure/flow conditions. Such measures were used to estimate aggregate valve leakage area described previously [8].

The Leonardo modified pulse duplicator originally adapted in 2011 with innovative features to measure PDVA from backlit valves remains in current use with improved spatial and temporal resolution capability (i.e., 0.001 centimeter^2^ and 1 microsecond). Valves tested in this system were recently compared to numerical simulations for contemporary porcine tissue and bovine pericardial bioprosthetic heart-valves and also with data from a similar independent pulse duplicator with excellent conformity [3].

A visco-elastic impedance adapter (VIA) models bulk ventricular visco-elasticity to produce physiological isovolumetric functionality has been utilized in several previous studies [3, 6-10]. Jennings et al. [13] in their use of the Leeds in vitro valve tester to study stentless porcine valves found VIA advantageous in ameliorating unphysiological left-ventricular pressure oscillations. Ventricular visco-elasticity associates with valve opening and closure dynamics, ventricular pressure rates (*dp/dt*), propensity for microbubbles, high intensity trancranial signals (HITS), cavitation, acoustic emissions, and vortex flow formations. Recently, Lee et al. [3] characterized VIA as a three-element Windkessel and utilized in fluid-structure interaction models of bioprosthetic heart valve dynamics. These models were validated with an experimental pulse duplicator.

As previously described, closed valve leakage was accurately measured in a quasi-steady pressure/flow apparatus calibrated with known small orifice areas [6-9]. Total valve leakage area was then edited into the experimental data and subsequently analyzed (leakage areas: SJM=0.0178 centimeter^2^; BV3D=0.003 centimeter^2^; Edwards Perimount™ Bioprosthesis=0.012 centimeter^2^).

### 2.2 Leonardo

Since only fleeting glimpses of prosthetic valve motion can be seen by the unaided eye, we developed a simple technique to quantify high-speed valve motion detail (Basic Leonardo, Figure 1). Described previously, this approach provides important analog data about valve motion [3, 5-11].

Figure 3 shows SJM and mTAVR are more similar, in the aortic position, with maximum TPs at or near 1.0. The rapid prototype BV3D and Edwards-Pericardial (control tissue valve) are more similar with maximum TPs near 0.25.

**Figure 3:**
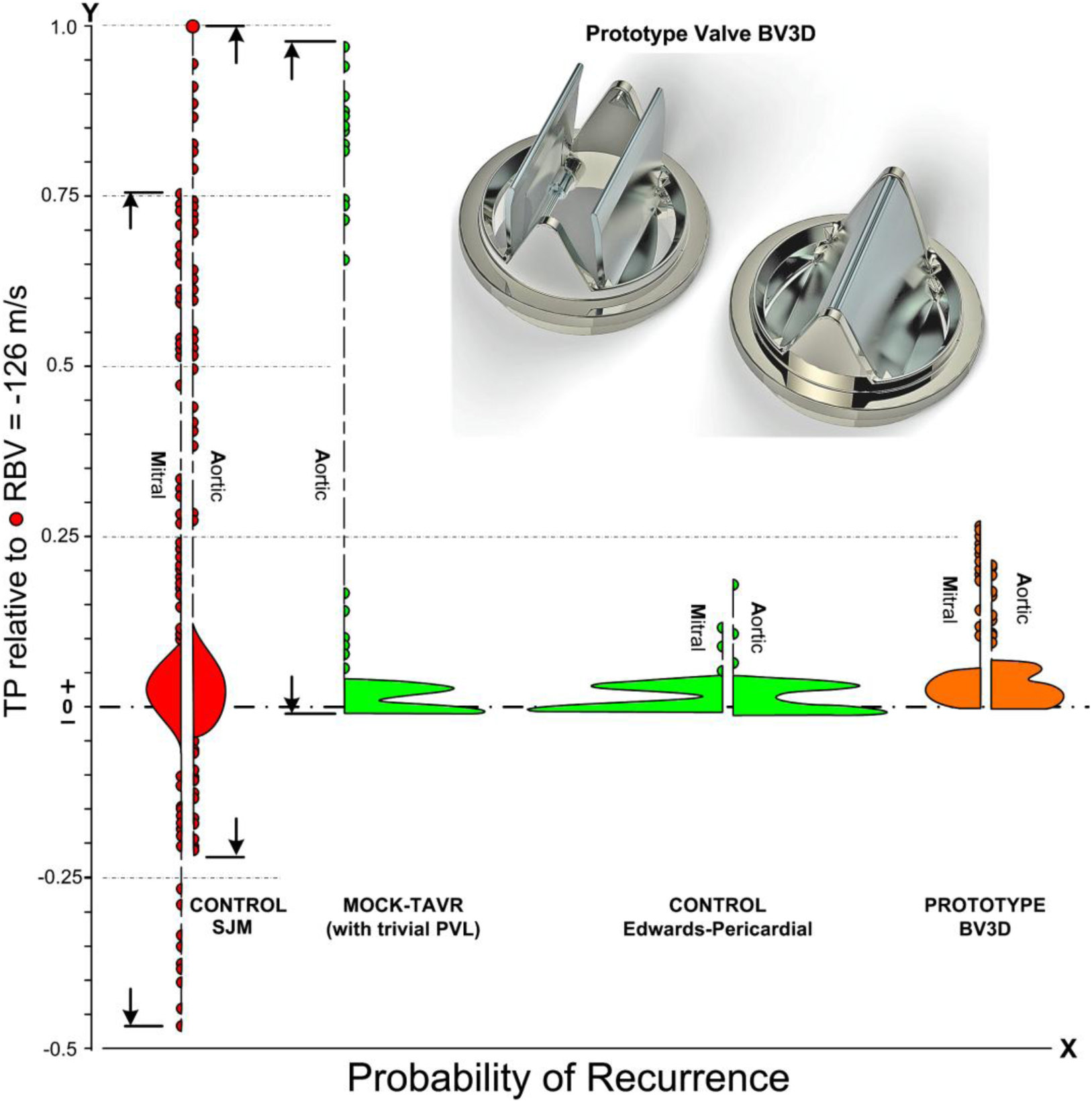
Dimensionless thrombogenic potential (TP) of valves (based on near closure RFV relative to −126 m/s). Early prototype valve rapid printed prototype bileaflet mechanical valve (BV3D) is shown in full open and closed views; Negative TPs involve water hammer and occluder rebound phenomena except for valve BV3D where further development is pending. Vioplot R* software analyzed a 10,500 dataset to create full-violin plots and Visio** software facilitated the split-violin graphics shown.

Vioplot R* software package and Visio 2002 ** have been utilized to create the split-violin plots used in this study. *R Core Team (2019) R: A Language and environment for statistical computing. R Foundation for Statistical computing, Vienna, Austria, URL https://www.R-project.org

** Microsoft, Redmond, WA, USA

### 2.3 Violin Plots and Statistical Synergies

The statistical method for visualization of the datasets utilized Violin Plots. Introduced in 1998 by Hinze and Nelsen [12], such plots attracted our attention as a powerful synergistic technique, to present voluminous quantitative or qualitative datasets. The violin plot is the union of density shape, kernel density estimate, and summary statistics relevant to boxplots. Figure 2 depicts TP probability density near valve closure and reveals different peaks, their position, and relative amplitudes. Readers will appreciate that broader split-violin regions convey more probable flow velocities than those in narrower regions. TPs for valves during the decelerating forward flow phase on the verge of closure were determined as RFVs relative to a control mechanical valve RFV value of −126 m/s. RFV was derived using time-dependent volumetric flow rate/PDVA.

**Figure 2:**
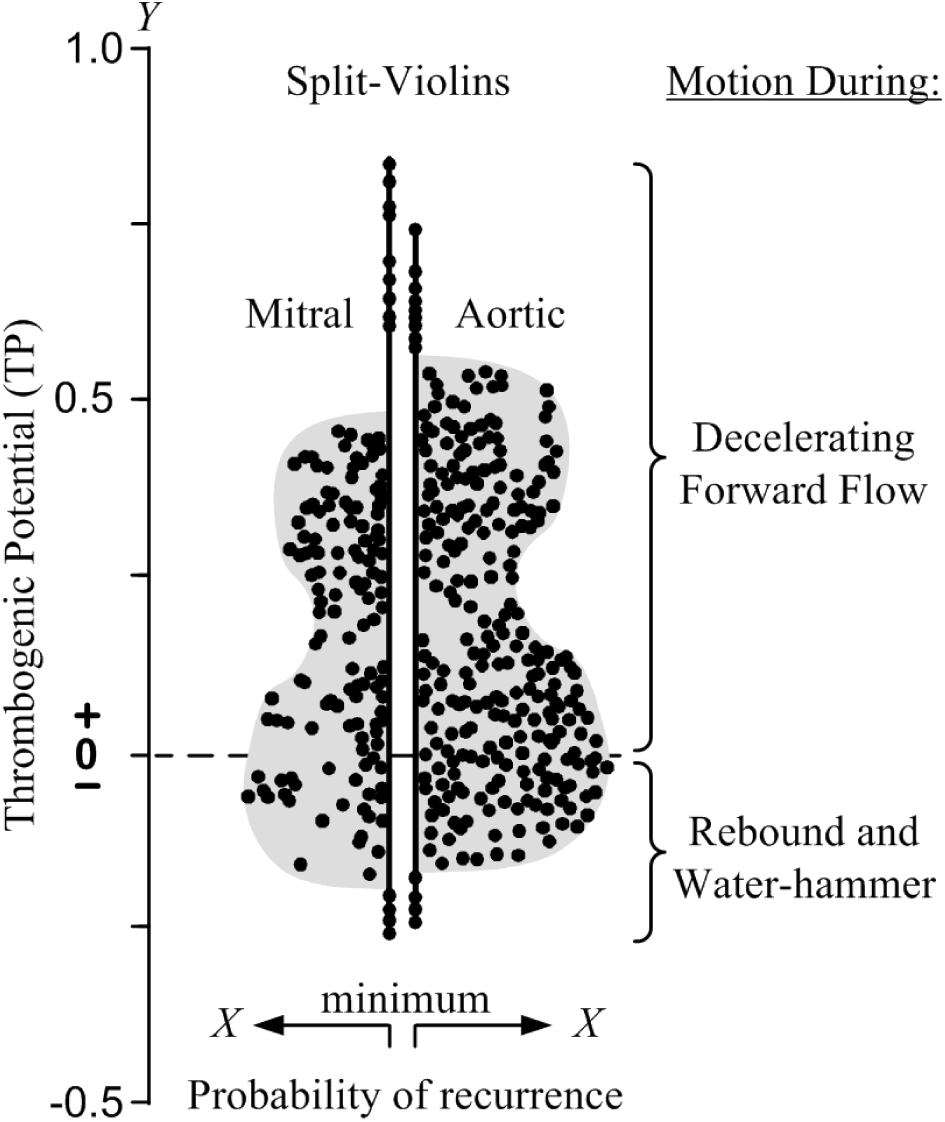
Sample split violin-plot visualization of inferred dimensionless valve thrombogenic potential (TP). Shaded profiles represent probability of recurrence of TP relative to RFV of −126 m/s). Full violin plots are symmetric and can be split, without loss of information, with each configuration representing independent data (n=150). Minimum X data values comprise those with minimum probability of recurrence but exhibit largest TP magnitudes.

### 2.4 Regurgitant Flow Velocity and Shear Force

Regurgitant flow velocity (RFV) magnitudes shown in Figure 3 can be considered a shear force surrogate associated with valve TP. Since flow velocity and fluid shear are related through flow velocity gradient, *a proxy for valve thrombogenic potential may be inferred by flow velocity*. Valve RFV is equal to volumetric flow rate, divided by the PDVA (flow rate/PDVA) and is an estimate of the regional instantaneous spatial average. Although TP cannot be assumed to increase linearly with flow velocity it is considered proportional to the inferred regional flow velocity (m/s) and independent of valve fabrication materials. The regional flow velocity determined by Leonardo is an estimated spatial average quantity (m/s) and not a site specific quantity. Since flow velocity gradients cannot be obtained with Leonardo, neither can shear force per unit area values (dyne/centimeter^2^) from measured RBVs (m/s).

### 2.5 Derivations and PDVA Synchronization Adjustment

Analog signal bandwidth differences between PDVA and transvalvular volumetric flow rate were 0-337 KHz vs. 0-100 Hz, respectively. The PDVA signal preceded the other signals by −2.5 ms based on step signal response thus a phase adjustment between the PDVA and the other signals was required. Accordingly, post experiment, we shifted the acquired PDVA signal 1 data acquisition interval of −3.4 ms [6].

### 2.6 Violin Plots with Relative Flow Velocity Scaling

Over time, as alternate TP assessment metrics evolved, valve thrombogenic potential appears to remain consistent by multiple determinations. For example, a widely used clinical MHV (SJM, St Jude Medical) has consistently exhibited high TP compared with controls regardless of determination method [6-10]. In all examples, determinations utilized valve motion as a component metric for obtaining TP [6-10].

The photograph in Figure 4 is from our 2011 publication [6] which captures saline jetting through small clearances intrinsic to a closed SJM aortic valve driven by a transvalvular back pressure of 120 mmHg.

**Figure 4:**
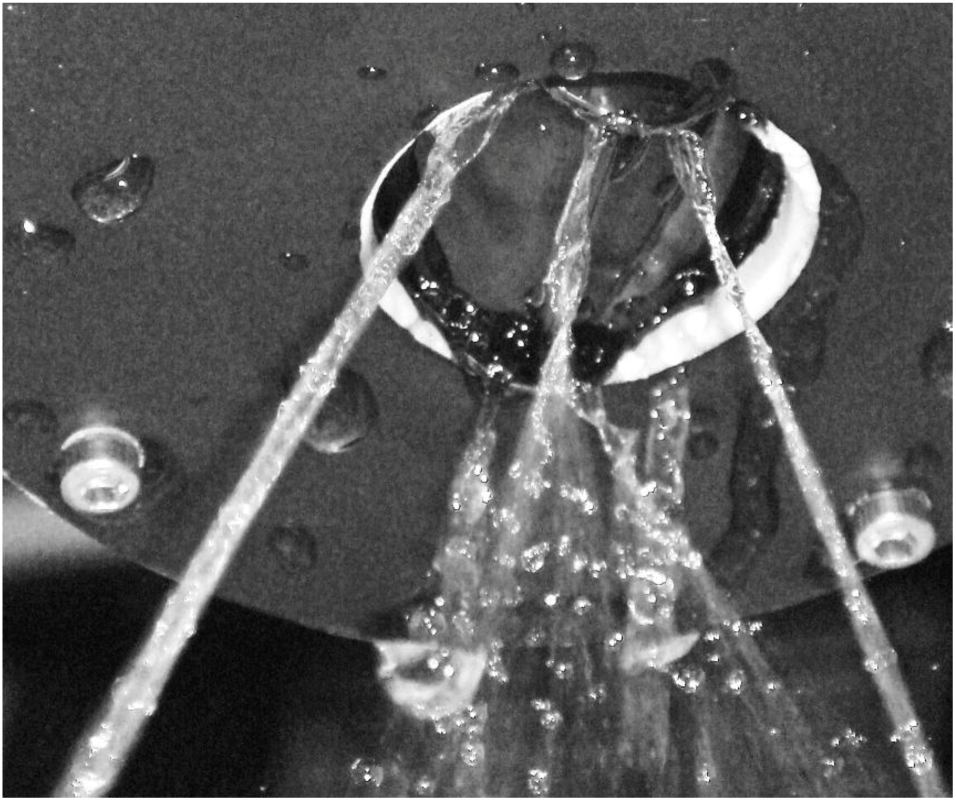
Photograph of multiple intra-valvular saline leakage jets passing through a closed St Jude Medical (SJM) mechanical valve. Transvalvular backpressure on closed valve is ca. 120 mmHg. Photo details: Camera SONY model DSC-T100, f/4.4, exposure time 1/50 s, and flash mode enabled.

Complex geometric gaps are present in all current mechanical valve designs. The total aggregate gap area for the closed SJM valve in Figure 4 is 0.0178 centimeter^2^ and under pulsatile flow conditions has a maximum spatially averaged regional flow velocity of ∼126 m/s near valve closure.

Readers are familiar with the qualitative echocardiographic signature of these retrograde jets during immediate post implant assessment of mechanical valves to detect peri-prosthetic leak; however, the magnitude and short duration of these flow transients cannot be resolved. In the absence of clinically significant hemolysis, thrombotic obstruction or thromboembolic events, the longer term implications of the conspicuous retrograde velocities, for recipient coagulation system-components, have remained largely unknown.

Recent clinical findings, that paravalvular leaks (PVL) are common (∼33%) after TAVR [14] identify micro-jets as a cause for hemolysis with thrombogenic consequences now evolving [15, 16]. Interestingly, lactate dehydrogenase (LDH) values have been largely ignored until recently when a reduction of 600 U/L was reported and considered significant [15]. LDH activity is expressed in U/L (International units) defined as the amount of enzyme that catalyzes the conversion of 1 micro-mol of substrate per minute under specified conditions. Evidence is emerging that the smaller the PVL, the higher the TP [8, 17].

PVL closure may have a therapeutic effect on the extent of hemolysis that some patients experience after valve surgery. Reduction of hemolysis is most likely in a patient with a mechanical valve and is not necessarily correlated with the volume of regurgitant flow or its subsequent reduction. Hemolysis associated with biological valves are less likely to respond to PVL closure suggesting that mechanisms other than net volume of regurgitation or micro-jets are present and should be considered [14, 15].

## 3. Results and Discussion

At near valve closure, flow dynamics can be considered analogous to transient valve stenosis, whereby regurgitation is increasingly constrained until complete, motionless, closed-valve conditions occur. During brief crucial moments preceding valve closure, localized prothrombotic microenvironments may be relevant to generation of high velocity leakage jets, flow unsteadiness, valve flutter, cyclic variability of PDVA, turbulence, and excessive shear forces that may induce blood element damage [15-31]. These influences may impact multiple valve types for:

- conduction abnormalities,
- reduced valve leaflet mobility, sub-clinical thrombosis [17, 23, 26-28, 30],
- potential for pannus formation,
- silent cerebral micro-infarction [24, 25],
- acute and sub-acute embolic stroke and other adverse cerebrovascular events including TIA [24, 26],
- cavitation and high intensity trans-cranial signals (HITS) [6].

Closure dynamics therefore may be directly related and analogous to:

1. Pro-thrombotic aspects of valve forward flow (e.g., valve stenosis) and arterial flows (e.g., arterial sclerosis).
2. Biomechanical and biochemical responses sufficient to exacerbate risk for pathologic thrombus formation and propagation.
3. Rebound and water-hammer for the SJM valve in the mitral position consistently manifested leaflet rebound observed as a momentary post closure partial re-opening driven by water-hammer power (transvalvular pressure × volumetric flow rate). This has been previously reported with magnified examples of high-resolution PDVA rebound data [7].
4. The SJM valve produces water-hammer with a maximum approaching −0.5; however, the mock-TAVR and Edwards-Pericardial do not. The highest probability of detection is as expected near 0.0 when bioprosthetic valves arrive at closure.

Figure 3 compares several contemporary and prototype cardiac valves near closure extending over ∼495 ms. One hundred fifty (150) RFV data points were accumulated for each valve over 10 consecutive cycles. The time increment was ∼3.4 ms which was the interval used to collect data over the total cycle time of 867 ms (cycle rate = 70 per minute). Inferred dimensionless TP of the test valves (Y axis) is referenced to an RFV of 126 m/s, the peak observed for the SJM MHV.

### 3.1 Study outcomes (previous and present reports) [6-10]

1. High amplitude RFVs and resultant supra-physiologic shear forces originating in small intra-and PVL gaps are mechanistic initiators of the clotting cascade.
2. Platelet activation is induced by high shear even with short exposure time [19, 20].
3. Closing dynamics disparity between MHV and bioprosthetic valves is chronically overlooked as a primary indicator of valve TP.
4. Current MHVs have overt RFV transients at closure relating to occluder non-response to flow deceleration and residual PDVA.
5. A mock-TAVR valve with trivial PVL generated high amplitude short duration RFVs.
6. For a given volumetric backflow rate near valve closure, the smaller the total residual leakage area, the greater the RFV magnitude.
7. The highest recorded RFVs were generated by the SJM valve compared to the tissue control valves (Edwards pericardial).
8. A spatial average of RFVs in immediate proximity to prototype valves may be a practical indicator in qualitative screening for valve TP.
9. Valve BV3D test data suggest that specific MHV leaflet geometries generate a closure force, during forward flow deceleration and prior to flow reversal, a potentially beneficial “soft closure” response.
10. Our results infer shear damage to formed blood elements and a resultant continuous thrombogenic response. This constitutes a mechanistic explanation for observed thrombogenic disparity, in prosthetic valve types and designs, now broadened to include observations of TAVR related thromboembolic events.
11. For MHVs, cyclic RFVs appear to be related to a prothrombotic state requiring chronic anti-coagulation.
12. Bioprosthetic valves appear not to generate a pro-thrombotic closing phase; however, TAVR prostheses with trivial PVL may have pathologic RFVs similar to those observed in MHVs [8].

### 3.2 Study limitations

Valve regional flow velocities are useful surrogates for flow induced blood cell damage potential; however, TP cannot be directly inferred from derived maximum flow velocities, considering the complexity of blood coagulation factors, recently reviewed by Rana and associates [21]. For example, the difference between laminar and turbulent flow, and their respective impact on blood component damage, awaits experimental evidence to validate numerical simulations [22]. In contrast to laminar flow, turbulent fluid flow is characterized by chaotic flow/pressure fluctuations with sufficient shear force to activate blood clotting mechanisms and influence von Willebrand factor (vWF) mechanisms [31].

## 4. Conclusions

Our prior work found valve motion and flow velocity constitute a primary contribution to valve TP and that closure-related functional deficit is a common shortcoming of current MHVs [6-9]. Progress, beyond the limitations of current prosthetic valves, requires appreciation of valve closure as a crucial factor. Very high-flow velocity short-duration RFVs near valve closure (Figure 4) result in transient blood cell damage each cardiac cycle and initiation of the coagulation cascade [20, 21]. Additionally, results suggest that designing a MHV with reduced TP, comparable to clinically well established tissue valves, is achievable with current technology. On a cautionary note, device manufacturers preoccupied with modifications of catheter delivered valves, to minimize residual leak, assume that a minor (trivial) leak reduces patient risk. However, we consistently observe smaller volume metric PVLs associate with higher magnitude RFVs and conversely; larger PVLs result in lower RFVs and shear forces. This is consistent with recently reported clinical experience where independent predictors of late leaflet thrombosis, up to 3 years post implant, were male sex *and PVL less than mild* [17]. Perhaps the most consequential outcome of our recent valve research is the use of data visualization, in the form of an exploratory graphic technique, to compare and screen valves, for inferred thrombotic potential, with well established clinical controls.

## Data Availability

All data relevant to the study are included in the article or uploaded as supplementary information.

## Footnotes

### Contributions

(I) Conception and design: LN Scotten; (II) Administrative support: LN Scotten; (III) Provision of study materials: LN Scotten; (IV) Collection and assembly of data: LN Scotten; (V) Data analysis and interpretation: LN Scotten, D Blundon; (VI) Manuscript writing: All authors; (VII) All authors have approved the final submitted version of the manuscript.

## Acknowledgements

Jason Nicholls utilized SolidWorks software for modeling and Keyshot software for rendering the valve prototype BV3D in central picture and Figure 3.

## Provenance and peer review

Not commissioned; externally peer reviewed

## Data sharing statement

All data relevant to the study are included in the article.

## Open access statement if applicable

Anyone can share, reuse, remix, or adapt this material, providing this is not done for commercial purposes and the original authors are credited and cited.

## Abbreviations and Acronyms (minimized)

BHV: bioprosthetic heart valve
BV3D: rapid prototype bileaflet mechanical valve (3D printed)
HITS: high intensity trancranial signal
ml/s: milliliters per second
m/s: meters per second
ms: milliseconds
mTAVR: mock-transcatheter aortic valve replacement
MHV: mechanical heart valve
PDVA: projected dynamic valve area
PVL: paravalvular leakage
RFV: regurgitant flow velocity
s: second
SJM: St Jude Medical
TP: thrombogenic potential
VIA: viscoelastic impedance adapter

